# Affect regulation in the context of sexual and gender minority stress: A scoping review protocol

**DOI:** 10.1101/2025.08.15.25332741

**Authors:** Daphne Y. Liu, Benjamin A. Swerdlow, Shao Yuan Chong, Nadia Kako, Alex Rubin, Nicholas S. Perry

## Abstract

**Objective:** To provide a broad, comprehensive picture of affect regulation in the context of sexual and gender minority stress, this scoping review aims to identify and synthesize methods, methodologies, and available evidence pertinent to emotion regulation and coping in the context of minority stress among sexual and gender minority (SGM) people.

**Introduction:** SGM people face disproportionately high rates of mental health problems due to experiences of minority stress and lack of social safety. Theories and growing evidence suggest that affect regulation plays a critical role in SGM people’s well-being in the face of minority stress. Researchers have largely studied emotion regulation, coping, and minority stress in distinct literatures, signaling a critical need to synthesize evidence across these bodies of research.

Inclusion criteria

We will review empirical studies that (1) include sexual and gender minority people, (2) measured at least one affect regulation construct, and (3) studied affect regulation in the context of sexual or gender minority stress.

**Methods:** Published and unpublished (i.e., grey literature) empirical studies written in English (no restrictions on publication date) will be searched using the following databases: PsycINFO (via EBSCO), Web of Science Core Collection (via Clarivate), PubMed (via National Library of Medicine), Gender Studies Database (via EBSCO), Sociological Abstracts (via ProQuest), and SocIndex with Full Text (via EBSCO). Grey literature will be additionally identified through searching on additional repositories and databases and emailing listservs of relevant organizations. Potentially relevant papers will first be screened based on title and abstract, followed by full-text screening, against inclusion criteria by two independent reviewers. Data on study characteristics and findings relevant to the review will be extracted by two independent reviewers. Frequency data relevant to each research question will be presented in tabular format, followed by a narrative summary of main findings, research gaps, and areas for future research.

---

Sexual and gender minority (SGM) people—individuals who do not identify as heterosexual or whose gender identity does not align with their sex assigned at birth [1–3]— are at disproportionately high risk for mental health problems (e.g., depression, anxiety, suicide attempts) [4,5]. The elevated rates of psychological problems reported by SGM people can be partly attributed to experiences of discrimination, prejudice, and stigma based on their minoritized sexual and/or gender identities, or sexual and gender minority stress (hereafter *minority stress*) [6–10]. Minority stressors exist at both the distal level (i.e., external, objective stressors, such as discriminatory hiring practices or enacted social rejection) and the proximal level (i.e., internal, subjective stressors that are theorized to result from exposure to external stressors, such as internalized identity-related stigma or expectations of rejection) [7,8,11]. The lack of social safety (i.e., reliable social connections, inclusion, and protection) that SGM people experience in their social environments in the presence of minority stressors also is thought to negatively impact their mental health [12].

The root causes of minority stress undoubtedly exist at the societal and institutional levels and require systemic interventions [13]. Nonetheless, the adverse affective experiences SGM endure as a result of exposure to minority stressors can have long-term adverse impact on their physical and mental health, thus calling for effective and context-appropriate coping and emotion regulation by the individual [14,15]. *Coping* refers to the process that is initiated in response to stress, which involves “constantly changing cognitive and behavioral efforts to manage specific external and/or internal demands that are appraised as taxing or exceeding the resources of the person” (p. 141) [16]. *Emotion regulation* refers to “the processes by which individuals influence which emotions they have, when they have them, and how they experience and express these emotions” (p. 275) [17]. Although distinct in several important ways, coping and emotion regulation are both goal-directed processes that share significant conceptual and measurement similarities [18,19]. Thus, in line with others [18,19], we use *affect regulation* as an umbrella term to refer to both coping and emotion regulation.

Growing theory and evidence suggest that SGM people’s abilities and behaviors to cope with and manage their affective experiences in the face of minority stress have critical implications for their psychological well-being [15,20–22]. For example, Meyer’s [8] minority stress model posits that individuals’ coping capacities can serve as an important moderator of the link between minority stress and mental health outcomes. Specifically, having access to and the awareness to use personal (e.g., self-acceptance) and group (e.g., an LGBTQ+ affirming church) coping resources may ameliorate and counteract the impact of minority stress on well-being. Additionally, certain proximal stressors, such as identity concealment, can potentially serve affect regulatory functions themselves in connection with anticipated possible distal stressors (e.g., by reducing the threat of antigay violence; Meyer, 2003), although use of these strategies may result in negative psychological and physical outcomes in the long run [20,23,24]. Hatzenbuehler’s [11] psychological mediation framework presents an alternative view of the role of affect regulation in which coping and emotion regulation are general psychological processes that serve as mediating mechanisms through which minority stress contributes to health disparities. As discussed by Hatzenbuehler [11], exposure to minority stress not only prompts affect regulation efforts, but also poses substantial challenges for affect regulation (e.g., insofar as it may be resource-depleting or furnish relatively few affordances that could facilitate regulation). Further, repeated exposure to minority stress may have deleterious consequences for the ongoing development of affect regulation skills and habits. Thus, over time, exposure to minority stress may jeopardize one’s affect regulation resources and abilities, thereby increasing risk for mental health problems. There is increasing support for the theory that minority stress increases maladaptive affect regulation processes (e.g., rumination), which in turn contributes to psychopathology (e.g., depressive symptoms) [25,26]. Together, both theories and growing empirical research support the important role of affect regulation in SGM people’s well-being in the face of minority stress. In fact, many psychological interventions tailored for SGM populations focus on enhancing affect regulation [27–30].

One barrier to advancing our understanding of affect regulation in the context of minority stress is that coping and emotion regulation represent largely distinct literatures. Put differently—and notwithstanding acknowledgements of the contributions of the stress and coping literatures to research on emotion regulation [31,32]—coping and emotion regulation have largely advanced as separate areas of research, with each having their own theoretical boundaries and frameworks [32,33]. For example, literature reviews will often focus on either the coping literature *or* the emotion regulation literature [34,35]. In parallel with the development of the coping and emotion regulation literatures, separate lines of empirical research have been conducted to understand coping [36] and emotion regulation [37] in the minority stress context, despite the fact that coping and emotion regulation have sometimes been treated as largely interchangeable in theoretical work on minority stress [11]). Given the substantial overlap between coping and emotion regulation and growing support for their relevance to the minority stress context and SGM people’s well-being, we argue that it is time to comprehensively examine the body of work that has focused on affect regulation in the minority stress context among SGM people.

As outlined above, minority stress theories and research have often drawn on constructs related to affect regulation. However, most affect regulation constructs that have been studied have not been developed for measuring regulation specifically in the context of minority stress context or based on SGM people’s lived experiences. Instead, researchers interested in studying affect regulation in the context of minority stress have commonly used more generalized affect regulation constructs and measures and applied them to minority stress [38–40]. On one hand, coping and emotion regulation are general psychological processes that are applicable to many situations, contexts, and populations and have far-ranging implications for psychological well-being [15,41,42]. Thus, using the frameworks and measures from the general coping and emotion regulation literatures may very well be appropriate. On the other hand, because minority stress represents a unique form of social stress that is salient to SGM people [20], it is unclear whether these general frameworks and measures fully capture the scope, nuances, and salient aspects of affect regulation SGM people engage in in response to minority stress (e.g., with respect to motives, tactics, etc.). Because of this, we think it is critical to evaluate the methodologies and methods researchers have used to assess affect regulation in the context of minority stress, as well as the extent to which researchers have taken a more bottom-up approach to studying affect regulation in the minority stress context based on SGM people’s lived experiences.

Taken together, we propose to conduct a scoping review to provide a broad, comprehensive picture of affect regulation in the minority stress context. The **objective** of this scoping review is to identify and synthesize methods, methodologies, and available evidence pertinent to affect regulation in the context of minority stress among SGM people. In order for a study to have examined “affect regulation in the context of minority stress”, the study needs to have measured affect regulation that takes place specifically in response to experienced or anticipated minority stress, experimentally induced minority stress when studying affect regulation, *or* have examined the interplay between affect regulation and minority stress, such as in moderation or mediation analyses (see Inclusion Criteria below for more details of the boundaries of this definition and examples that fall within and outside the scope of this review). We chose to conduct a scoping review because it is particularly well-suited for comprehensively mapping research evidence on a topic, synthesizing broad literatures that have not been integrated, and providing an understanding of the state of the literature and identify gaps [43,44]. A preliminary search of PsycINFO, PubMed, Google Scholar, the Cochrane Database of Systematic Reviews and JBI Evidence Synthesis was conducted, and no published or in-progress systematic reviews or scoping reviews on this topic were identified. A comprehensive review of the extant literature on affect regulation can not only provide a clear picture of how researchers have studied affect regulation in the minority stress context, but also identify important research gaps and future directions. This review is intended to encourage dialogue and collaboration among researchers across disciplines and guide future interdisciplinary collaborations. We also consider this scoping review to be an initial step for determining whether systematic reviews on certain topics within this broader integration are warranted.

## Review Questions

This scoping review aims to address the following four research questions:

1. What kinds of research questions have researchers tried to answer when investigating affect regulation in the context of minority stress?
2. What research designs (e.g., cross-sectional or longitudinal; quantitative, qualitative, or mixed methods; experimental, quasi-experimental, or descriptive) and analyses (e.g., mediation, moderation, thematic analysis) have researchers used to address these questions?
3. How have researchers measured affect regulation in the context of minority stress (e.g., global self-report surveys, laboratory-based observations, daily diaries, ecological momentary assessment, qualitative interviews)?
4. To what extent have researchers attended to the experiences of subgroups among SGM populations or to intersecting identities of SGM people (e.g., SGM people of color)?

## Materials and Methods

This scoping review protocol was developed and written according to guidelines proposed by the Joanna Briggs Institute (JBI) Methodology Group [45]. We included a completed checklist of preferred reporting items recommended for protocol reviews (see S1 Table) [46], which was adapted from the preferred reporting items for systematic review and meta-analysis protocols (PRISMA-P) [47]. This scoping review will be conducted in accordance with the *a priori* protocol developed based on JBI methodology for scoping reviews [44,48]. This protocol has been pre-registered on Open Science Framework (OSF; https://osf.io/k456p/?view_only=5d4f9a7b5cec42d880aad16cadc96a22). The results of this scoping review will be reported according the Preferred Reporting Items for Systematic Reviews and Meta-Analysis: Extension for Scoping Reviews (PRISMA-ScR; Tricco et al., 2018). Any deviations from the protocol will be reported in the Method section of the final publication following the completion of this review. This scoping review involves reviewing published and grey literature and thus does not require Institutional Review Board review and approval. We will not have access to data that could identify participants of reviewed studies.

## Inclusion Criteria

The inclusion criteria of this scoping review are organized based on JBI’s Population, Concept, and Context (PCC) framework [45].

### Participants

#### All, or a subsample, of participants of the included studies need to have identified themselves as SGM

Participants may be of any age, and there is no specified minimum number or percent of the subsample that needs to have identified as SGM. However, studies are excluded if no participants identified as SGM, or if the authors did not report participants’ sexual or gender identity.

#### Concept

The study needs to have assessed **at least one affect regulation construct**. Affect regulation encompasses emotion regulation and coping. The affect regulation construct may be related to one’s general affect regulation ability or their frequency or tendency to engage in a specific affect regulation strategy or behavior. The study may have assessed affect regulation or coping using any method, and data may be quantitative or qualitative. In order for the study to meet the Concept inclusion criteria, the study methods need to have included some direct assessment of affect regulation (i.e., methodologically). Some examples of this include: (a) using one or more measures that are designed to assess people’s affect regulation beliefs, tendencies, abilities, or strategies; (b) explicitly asking participants how they regulated their affect in a particular instance; or (c) experimentally instructing participants to regulate their affect or observing how participants regulated affect in response to a stimulus.

It is worth noting that there are many different constructs related to affect regulation that have been studied in the literature, and there are many behaviors that may be conceptualized as having affect-regulating features (e.g., substance use). For the purposes of this scoping review, it must be clear from the paper that the construct that the authors assessed was explicitly measured *as an affect regulation strategy*; in other words, simply conceptualizing the construct as an affect regulation strategy (e.g., as described in the Introduction section of the publication) is not sufficient. For example, a paper that used the substance use subscale of the COPE Inventory (e.g., “I use alcohol or drugs to make myself feel better”) would likely qualify for this inclusion criterion, whereas a paper that used a measure of substance use frequency (“In the past week, how many times have you used tobacco?”) might not qualify even if the authors included some discussion of the possibility that substance use could be motivated by a desire to reduce stress. This is because a central feature of affect regulation is a goal-directed process that involves the activation of a goal to regulate affect and requires a person’s active efforts to manage internal or external demands or influence emotions [32]. This effort may come from oneself (e.g., a person suppresses their emotional expressions when anticipating discrimination) or from other people (e.g., a person’s spouse verbalizes affirmations to help the person cope with internalized stigma).

Though perceived affect regulation abilities and behaviors are acceptable constructs of affect regulation (e.g., subscale or total scores of Difficulties in Emotion Regulation of Scale) [50], perceived phenomena of one’s environment with no clear indication of any affect regulation goals or efforts do not meet inclusion criteria of concept (e.g., perceived availability of social support).

#### Context

Included studies need to have studied affect regulation **in the context of sexual or gender minority stress**. The spirit of this inclusion criterion is ensuring that we are including research that is at the *intersection* between affect regulation and minority stress, rather than studying these two constructs separately without directly connecting them. In line with this goal, we delineate several forms the study design can take that would meet the inclusion criteria of Context. The most straightforward case is when researchers included questions or scale items that explicitly asked participants about their affect regulation related to minority stress experiences (e.g., “Which of the following behaviors do you typically engage in to cope with experiences of discrimination related to your sexual orientation?”). A second case is when researchers experimentally induced a minority stress context in which affect regulation is assessed (e.g., exposing SGM participants to passages containing homophobic content and asking them to engage in cognitive reappraisal to improve their affect). Another way the study could have studied affect regulation in the context of minority stress is to directly examine affect regulation in their analyses relevant to minority stress, such as through mediation or moderation analyses. In the case of mediation, the authors could have examined whether rumination (an emotion regulation strategy) explains the association between recent experiences with gender-based discrimination and current depressive symptoms by testing the following mediation in line with the psychological mediation framework [15]: discrimination → rumination → depressive symptoms. In the case of moderation, the authors could have examined either how affect regulation moderates the link between minority stress and an outcome (e.g., test whether active coping styles attenuates the association between internalized transphobia and mental health) or how minority stress moderates the link between affect regulation and an outcome (e.g., test whether using alcohol to cope is more strongly associated with subsequent depression for those who experience more frequent discrimination).

On the other hand, a study should not be included if the researchers did not assess affect regulation specifically in response to experienced or anticipated minority stress, did not experimentally induce minority stress when studying affect regulation, *and* did not examine their interplay in mediation or moderation analyses. We describe two example scenarios in which the inclusion criteria of Context are *not* met. One scenario is where the authors simply examined the bivariate association between general affect regulation and minority stress with no clear indication that affect regulation takes place in response to minority stress (e.g., test whether those who more frequently use acceptance as an emotion regulation strategy tend to report lower internalized homophobia). A second scenario that does not meet this inclusion criteria is when a study examined how general affect regulation and minority stress constructs are separately associated with other construct(s) in the study, with no clear indication that affect regulation takes place in response to minority stress. For example, a study that examined how passive coping and identity concealment independently predict one’s depressive symptoms one month later by including these constructs simultaneously in a regression model (without examining their interaction with each other) would fall outside the scope of this review.

#### Types of Evidence Sources

This review will include published and unpublished research papers that feature empirical studies, defined as studies that derive scientific evidence based on concrete data collected from participants. We will include cross-sectional and longitudinal studies, studies that collect quantitative or qualitative data, and studies using a variety of methods for assessing affect regulation (e.g., surveys, laboratory-based experiments, experience sampling method, daily diaries, qualitative interviews). We will exclude books, book chapters, reviews, opinion papers, commentaries, and protocol papers. We will limit our review to only papers written in English given that all authors are trained in conducting research in English.

### Search Strategy

The search strategy will aim to locate published and unpublished studies that meet our inclusion criteria. This strategy was developed collectively by the authors and in consultation with an experienced research librarian at the University of Denver. The following databases will be used to search for relevant papers: PsycINFO (via EBSCO), Web of Science Core Collection (via Clarivate), PubMed (via National Library of Medicine), Gender Studies Database (via EBSCO), Sociological Abstracts (via ProQuest), SocIndex with Full Text (via EBSCO). Grey literature will be additionally searched via the following methods: (a) Google Scholar, (b) commonly used social sciences repositories (i.e., PsyArXiv, PsychArchives, OSF preprints, Social Science Research Network[SSRN]), (c) dissertation and thesis databases (i.e., ProQuest Dissertations & Theses Global, Open Access Theses and Dissertations WorldCat), and (d) emails soliciting unpublished research through relevant organizations’ listservs (i.e., Society for Affective Science, American Psychological Association Division 44: Society for the Psychology of Sexual Orientation and Gender Diversity, Association for Association for Behavioural and Cognitive Therapies).

A list of search terms pertaining to inclusion criteria was formulated to search for relevant papers across databases. The search terms can be grouped into three categories: SGM populations (e.g., *LGBT\**, *sexual and gender minorit\**), affect regulation (e.g., *coping*, *emotion regulation*), and minority stress (e.g., *minority stress*, *discrimination*), which correspond to inclusion criteria of participants, concept, and context, respectively. When searching for relevant papers across databases, the three categories of search terms will be connected with AND (as all of them need to be present according to our inclusion criteria). The generation of the search terms was informed by authors’ and the consulting librarian’s collective content expertise in relevant literatures, prior scoping and systematic reviews on related topics, and publicly available search hedges (i.e., expert-developed combination of search terms on a topic area) [51,52]. For PsycINFO and PubMed, a list of subject headings specific to the respective databases was also used to identifying additional potentially relevant articles. See S2 Table for a complete list of search terms and subject headings used in our search, and S3 Table for full search strategy for PsycINFO.

### Study Selection

Following the search, all identified citations across databases will be collated and uploaded into Rayyan [53], a systematic review management tool. The titles and abstracts of these citations will be screened by two independent reviewers for assessment against the inclusion criteria for the review. Each reviewer may indicate whether the article meets the inclusion criteria by indicating “include”, “maybe”, or “exclude” (the reviewer may indicate “maybe” when there isn’t sufficient information in the title and abstract to determine its eligibility). Potentially relevant sources (i.e., those that are marked as “yes” or “maybe” during initial screening) will be retrieved in full and imported into Rayyan. During this initial screening process, in the case where one reviewer indicates “include” or “maybe” and the other reviewer indicates “exclude”, disagreements will be resolved through discussion between the two reviewers. If disagreements persist, disagreements will be discussed as a team, with a final decision made by the first author (DYL). In cases where one reviewer indicates “yes” and the other “maybe,” the citation will proceed to full-text screening without the need to resolve the discrepancy at this stage of the process.

The full text of selected citations will be assessed in detail against the inclusion criteria by two or more independent reviewers, each making an “include” or “exclude” decision based on whether the source meets the inclusion criteria. Reasons for exclusion during the full-text review stage will be recorded and reported in the scoping review. During this process, any disagreements between the two reviewers will be resolved through discussion between the two reviewers; any persisting disagreements will be discussed as a team, with a final decision made by the first author (DYL). The results of the search and the study inclusion process will be reported in full in the final scoping review and presented in a PRISMA flow diagram [54].

To facilitate training and consistency across authors, a pilot screening exercise will take place for both the initial abstract screening and the full-text screening phase. For each of these stages, 15 sources will be selected for pilot screening procedures and training screeners, and these sources will be screened by all members of the research team who carry out article screening tasks. The process of searching, screening, and selecting eligible sources as well as team discussions may reveal potentially relevant search terms and constructs and result in modifications of the search strategy (e.g., search terms, concepts) during the review process. Any modifications will be documented in the final publication of this review.

### Data Extraction

Data will be extracted from papers included in the scoping review by at least two independent reviewers. The data extracted will include specific details about the participants, concept, context, study methods relevant to the review questions (see S4 Table for a draft data extraction form). To test the feasibility of the planned data extraction process and facilitate training and consistency across the team, a pilot data extraction exercise will be carried out. Specifically, research team members who will participate in the data extraction will be paired, and each pair will be assigned one quantitative and one qualitative study for data extraction. They will then meet to compare their data extraction sheets. The team will then meet as a group to discuss the pilot data extraction exercise and its feasibility and decide on any necessary modifications and refinements to be made to the data extraction process.

Any modifications made during the data extraction process will be reported in the final publication of this review. When appropriate, authors of papers will be contacted to request missing or additional data. Similar to the screening stage, any disagreements on data extraction that arise between the reviewers will be resolved through discussion between the two reviewers; if disagreements persist, disagreements will be discussed as a team, with a final decision made by the first author (DYL). Formal assessment of study quality and risk of bias individual sources of evidence will not be performed as it is outside the scope of the scoping review [48].

### Data Analysis and Presentation

For each research question, we plan to conduct basic frequency analyses and present relevant descriptive data in tables. In line with the review objective and research questions, we plan to include tables summarizing (1) different types of research questions researchers have investigated relevant to affect regulation in the context of minority stress (e.g., grouping of different research questions based on themes and descriptive frequencies of the themes); (2) frequencies of different types of research designs and analytic approaches researchers have used in these studies (e.g., number of studies that utilized quantitative, qualitative, and mixed methods); (3) frequencies of different ways researchers have measured affect regulation in the minority stress context (e.g., number of studies that utilized global self-report surveys, daily diaries, qualitative interviews, etc.; number of studies using generic measures of affect regulation and measures specifically designed to address SGM experiences), and (4) the extent to which researchers explicitly attended to experiences of SGM subgroups and intersectionality of identities (i.e., grouping of different ways researchers addressed subgroup experiences and intersectionality based on themes, followed by descriptive frequencies). Presentation of data for each research question will be followed by a narrative summary of main findings and takeaways, as well as gaps in the literature [55].

### Study Timeline

At the time of registering this protocol on OSF (July 11, 2025), the first author (DYL) had completed literature search (conducted on June 19, 2025) from the following databases: PsycINFO (via EBSCO), Web of Science Core Collection (via Clarivate), PubMed (via National Library of Medicine), Gender Studies Database (via EBSCO), Sociological Abstracts (via ProQuest), SocIndex with Full Text (via EBSCO). Downloaded citations were then uploaded to Rayyan and duplicates were removed by the first author (completed on July 5, 2025). The scoping review research team (i.e., authors of this protocol and additional undergraduate-level and post-baccalaureate research assistants) met every weekly from May to July, 2025 to develop the protocol and go through training of the title and abstract screening. Title and abstract screening via Rayyan started on July 9, 2025.

We anticipate the next steps of this scoping review will take 18 months to complete.

Specifically, the following timeline associated with each stage is expected: (a) title and abstract screening (4 months), (b) full-text screening (4 months), (c) data extraction (4 months), (d) data organization and synthesis (2 months), and (c) manuscript writing (4 months). Planned search of the grey literature will take place for the next four months and all citations are expected to be screened by the time full-text screening stage is complete.

## Data Availability

No research data are reported in this submission.

## Supporting Information Captions

**S1 Table:** Completed checklist recommended items to address in a scoping review. Adapted from the PRISMA-P (Preferred Reporting Items for Systematic review and Meta-Analysis Protocols) 2015 checklist*.

| Section and topic | Item No | Checklist item | Page #/Location |
| --- | --- | --- | --- |
| **ADMINISTRATIVE INFORMATION** |  |  |  |
| Title: |  |  |  |
| Identification | 1a | Identify the report as a protocol of a scoping review | Study Title |
| Update | 1b | If the protocol is for an update of a previous scoping review, identify as such | N/A |
| Registration | 2 | If registered, provide the name of the registry (such as JBI) and registration number | p. 10 |
| Authors: |  |  |  |
| Contact | 3a | Provide name, institutional affiliation, email address of all protocol authors; provide physical mailing address of corresponding author | Title page |
| Contributions | 3b | Describe contributions of protocol authors and identify the guarantor of the review | Title page |
| Amendments | 4 | If the protocol represents an amendment of a previously completed or published protocol, identify as such and list changes; otherwise, state plan for documenting important protocol amendments | p. 10 |
| Support: |  |  |  |
| Sources | 5a | Indicate sources of financial or other support for the review | Title page |
| Sponsor | 5b | Provide name for the review funder and/or sponsor | N/A |
| Role of sponsor or funder | 5c | Describe roles of funder(s), sponsor(s), and/or institution(s), if any, in developing the protocol | N/A |
| **INTRODUCTION** |  |  |  |
| Rationale | 6 | Describe the rationale for the review in the context of what is already known (Note: Consider providing a rationale for the choice of conducting a scoping review as compared to other evidence synthesis approaches) | p. 5-9 |
| Objectives | 7 | Provide an explicit statement of the question(s) the review will address with reference to the inclusion/ exclusion criteria | p. 9 |
| **METHODS** |  |  |  |
| Eligibility criteria | 8 | Specify the study characteristics (such as PICO, study design, setting, timeframe) and report characteristics (such as years considered, language, publication status) to be used as criteria for eligibility for the review | p. 10-13 |
| Information sources | 9 | Describe all intended information sources (such as electronic databases, contact with study authors, trial registers or other grey literature sources) with planned dates of coverage | p. 14-15 |
| Search strategy | 10 | Present draft of search strategy to be used for at least one electronic database, including planned limits, such that it could be repeated | S3 Table |
| Study records: |  |  |  |
| Data management | 11a | Describe the mechanism(s) that will be used to manage records and data throughout the review | p. 15-16 |
| Selection process | 11b | State the process that will be used for selecting studies (such as two independent reviewers) through each phase of the review (that is, screening, eligibility and inclusion) | p. 15-16 |
| Data collection process | 11c | Describe planned method of extracting data from reports (such as piloting forms, done independently, in duplicate), any processes for obtaining and confirming data from investigators | p. 16-17 |
| Data items | 12 | List and define all variables for which data will be sought (such as PICO items, funding sources), any preplanned data assumptions and simplifications (Note: Scoping reviews may not use PICO and instead may use JBI's Population, Concept, and Context [PCC] or another approach to reporting eligibility criteria) | S4 Table |
| Outcomes and prioritization | 13 | List and define all outcomes for which data will be sought, including prioritization of main and additional outcomes, with rationale (Note: Scoping reviews may not extract outcome data, so this can refer to whichever data items are extracted) | S4 Table |
| Risk of bias in individual studies | 14 | If this is to occur, describe anticipated methods for assessing risk of bias of individual studies, including whether this will be done at the outcome or study level, or both; state how this information will be used in data synthesis (Note: Scoping reviews typically do not include risk of bias assessment, but this information should be described if it will occur) | N/A |
| Data synthesis | 15a | Describe criteria under which study data will be presented (Note: Scoping reviews do not typically include quantitative synthesis of study data, but should still describe in advance how extracted data are anticipated to be presented in the resulting review) | p. 17-18 |
|  | 15b | Describe the planned approach to how extracted data will be presented (such as figures, tables, evidence gaps maps) | p. 17-18 |
|  | 15c | Describe any proposed additional analyses (such as thematic analyses) (Note: The JBI methodological guidance does not recommend undertaking thematic analysis as this synthesis of data should ideally occur following methodological appraisal of the included sources) | N/A |
|  | 15d | If quantitative synthesis is not appropriate, describe the type of summary planned | p. 17-18 |
| Meta-bias(es) | 16 | Specify any planned assessment of meta-bias(es) (such as publication bias across studies, selective reporting within studies) (Note: Scoping reviews typically do not include assessment of meta-bias[es], but this information should be described if it will occur) | N/A |
| Confidence in cumulative evidence | 17 | Describe how the strength of the body of evidence will be assessed (such as GRADE) | N/A |

\*This checklist is adapted by Peters and colleagues [1] based on the PRISMA-P (Preferred Reporting Items for Systematic review and Meta-Analysis Protocols) 2015 checklist [2]

---

**S2 Table:** Complete list of search terms and subject headings.

|  | **SGM Populations** | **Affect Regulation** | **Minority Stress** |
| --- | --- | --- | --- |
| **Search Terms** | LGBT* OR GLBT* OR “sexual and gender minorit*” OR “sexual gender minorit*” OR “sexual minorit*” OR “sexually diverse” OR “sexual diverse” OR gay* OR lesbian* OR homosexual* OR “same-sex” OR “non-heterosexual” OR “same-gender” OR bisexual* OR “bi-sexual*” OR pansexual* OR polysexual* OR demisexual OR asexual* OR queer OR “gender minorit*” OR “non-cisgender*” OR genderqueer OR “gender-queer” OR nonbinary OR “non-binary” OR agender* OR “gender-expansive” OR “gender divers*” OR genderfluid* OR “gender-fluid*” OR “two-spirit*” OR “bigender” OR “intersex” OR “gender non-confirming” OR “gender nonconfirming” OR “gender variant” OR “third gender*” OR transgender* OR transfeminine OR transmasculine OR transsexual* OR “trans man” OR “trans men” OR “trans woman” OR “trans women” OR “trans person*” OR “trans people” OR “trans youth” OR “trans adolescent*” OR “trans boy*” OR “trans girl*” OR “trans child*” OR “men who have sex with” OR “males who have sex with” OR “women who have sex with” OR “females who have sex with” | cope OR coping OR coped OR “emotion regulat*” OR “emotion regulatory” OR “emotion dysregulation” OR “regulate emotion*” OR “regulating emotion*” OR “regulated emotion*” OR “affect regulat*” OR “affect regulatory” OR “affect dysregulation” OR “regulate affect*” OR “regulating affect*” OR “regulated affect*” OR “emotion management” OR “manage emotion*” OR “managing emotion*” OR “mood regulation” OR “mood regulatory” OR “regulat* mood” OR “stress regulation” OR “regulate stress*” OR “regulating stress*” OR “regulated stress*” OR “stress management” OR “manage stress*” OR “managing stress*” OR “managed stress*” OR “cognitive reappraisal” OR “expressive suppression” OR “emotion* suppression” OR ruminati* OR distraction | **Search Terms:** “minority stress*” OR discriminat* OR violence OR mistreat* OR oppress* OR marginalize* OR microaggress* OR victimi* OR reject* OR stigma* OR prejudic* OR heterosexis* OR homonegativ* OR harass* OR homophobi* OR transphobi* OR queerphobic*, biphobi*, OR cisgenderis* OR “cis-genderis*” OR cisheterosexis* OR “cis-heterosexis*” OR cisnormativ* OR “cis-normativ*” OR heteronormativ* OR “hetero-normativ*” OR “hypervigilan*” OR “rejection sensitiv*” OR “gender dysphori*” OR “anti-LGBT*” |
| **PsycINFO Subject Headings** | **PsycINFO subject headings:** DE “LGBTQ” OR DE “LGBTQ Rights” OR DE “LGBTQ Parents” OR DE “Asexuality” OR DE “Bisexuality” OR DE “Gender Nonbinary” OR DE “Gender Nonconforming” OR DE “Intersex” OR DE “Lesbianism” OR DE “Transgender” OR DE “Transgender (Attitudes Toward)” OR DE “Homosexuality” OR DE “Homosexuality (Attitudes Toward)” OR DE “Male | **PsycINFO subject headings:** DE “Affect Regulation” OR DE “Emotion Regulation” OR DE “Emotional Regulation” OR DE “Stress and Coping Measures” OR DE “Coping Behavior” OR DE “Coping Style” OR DE “Stress Management” OR DE “Expressive Suppression” OR DE “Emotional Control” OR DE “Anger Control” OR DE “Self- | **PsycINFO subject headings:** DE “Minority Stress” OR DE “Marginalization” OR DE “Prejudice” OR DE “Stigma” OR DE “Victimization” OR DE “Discrimination” OR DE “Sex Discrimination” OR DE “Social Discrimination” OR DE “Microaggression” OR DE “Gender Dysphoria” OR DE “Harassment” OR DE |
|  | Homosexuality” OR DE “Female Homosexuality” OR DE “Sexual Minority Groups” OR DE “Gender Identity” OR DE “Sexual Identity” OR DE “Pansexuality” OR DE “Same Sex Marriage” OR DE “Same Sex Couples” OR DE “Transsexualism” | Regulation” OR DE “Cognitive Appraisal” OR DE “Cognitive Control” OR DE “Thought Suppression” OR DE “Suppression (Defense Mechanism)” | “Hate Crimes” OR DE “Hate” OR DE “Aggressive Behavior” OR DE “Oppression” |
| **PubMed**<br>**MeSH**<br>**Terms:** | “Sexual and Gender Minorities”[Mesh:NoExp] OR “Bisexuality”[Mesh:NoExp] OR “Homosexuality”[Mesh:NoExp] OR “Homosexuality, Female”[Mesh:NoExp] OR “Homosexuality, Male”[Mesh:NoExp] OR “Transgender Persons”[Mesh:NoExp] OR “Intersex Persons”[Mesh:NoExp] OR “Transsexualism”[Mesh:NoExp] OR “Bigender”[Mesh:NoExp] OR “Gender-Nonconforming Persons”[Mesh:NoExp] | “Emotional Regulation”[Mesh:NoExp] OR “Coping Skills”[Mesh:NoExp] OR “Self-Control”[Mesh:NoExp] | “Social Stigma”[Mesh:NoExp] OR “Social Discrimination”[Mesh:NoExp] OR “Perceived Distrimination”[Mesh:NoExp] OR “Violence”[Mesh:NoExp] OR “Social Marginalization”[Mesh:NoExp] OR “Gender Dysphoria”[Mesh:NoExp] OR “Prejudice”[Mesh:NoExp] OR “Aggression”[Mesh:NoExp] OR “Microaggression”[Mesh:NoExp] OR "Rejection, Psychology"[Mesh:NoExp] OR “Harassment, Non-Sexual”[Mesh:NoExp] |

*Note:* Because PubMed only allows truncation used for search terms that have four or more characters we used “gay” OR “gays” instead of “gay\*” in the search term for PubMed. We did not include the following specific affect regulation strategies in our affect regulation search terms because including these would explode the search and result in mostly irrelevant search results: problem solving, acceptance, avoidance.

**S3 Table:** Full search strategy for PsycINFO (via EBSCO)

|  |  |
| --- | --- |
| 1. | LGBT* OR GLBT* OR “sexual and gender minorit*” OR “sexual gender minorit*” OR “sexual minorit*” OR “sexually diverse” OR “sexual diverse” OR gay* OR lesbian* OR homosexual* OR “same-sex” OR “non-heterosexual” OR “same-gender” OR bisexual* OR “bi-sexual*” OR pansexual* OR polysexual* OR demisexual OR asexual* OR queer OR “gender minorit*” OR “non-cisgender*” OR genderqueer OR “gender-queer” OR nonbinary OR “non-binary” OR agender* OR “gender-expansive” OR “gender divers*” OR genderfluid* OR “gender-fluid*” OR “two-spirit*” OR “bigender” OR “intersex” OR “gender non-confirming” OR “gender nonconfirming” OR “gender variant” OR “third gender*” OR transgender* OR transfeminine OR transmasculine OR transsexual* OR “trans man” OR “trans men” OR “trans woman” OR “trans women” OR “trans person*” OR “trans people” OR “trans youth” OR “trans adolescent*” OR “trans boy*” OR “trans girl*” OR “trans child*” OR “men who have sex with” OR “males who have sex with” OR “women who have sex with” OR “females who have sex with” OR DE “LGBTQ” OR DE “LGBTQ Rights” OR DE “LGBTQ Parents” OR DE “Asexuality” OR DE “Bisexuality” OR DE “Gender Nonbinary” OR DE “Gender Nonconforming” OR DE “Intersex” OR DE “Lesbianism” OR DE “Transgender” |
| 2. | DE “LGBTQ” OR DE “LGBTQ Rights” OR DE “LGBTQ Parents” OR DE “Asexuality” OR DE “Bisexuality” OR DE “Gender Nonbinary” OR DE “Gender Nonconforming” OR DE “Intersex” OR DE “Lesbianism” OR DE “Transgender” OR DE “Transgender (Attitudes Toward)” OR DE “Homosexuality” OR DE “Homosexuality (Attitudes Toward)” OR DE “Male Homosexuality” OR DE “Female Homosexuality” OR DE “Sexual Minority Groups” OR DE “Gender Identity” OR DE “Sexual Identity” OR DE “Pansexuality” OR DE “Same Sex Marriage” OR DE “Same Sex Couples” OR DE “Transsexualism” |
| 3. | S1 OR S2 |
| 4. | cope OR coping OR coped OR “emotion regulat*” OR “emotion regulatory” OR “emotion dysregulation” OR “regulate emotion*” OR “regulating emotion*” OR “regulated emotion*” OR “affect regulat*” OR “affect regulatory” OR “affect dysregulation” OR “regulate affect*” OR “regulating affect*” OR “regulated affect*” OR “emotion management” OR “manage emotion*” OR “managing emotion*” OR “mood regulation” OR “mood regulatory” OR “regulat* mood” OR “stress regulation” OR “regulate stress*” OR “regulating stress*” OR “regulated stress*” OR “stress management” OR “manage stress*” OR “managing stress*” OR “managed stress*” OR “cognitive reappraisal” OR “expressive suppression” OR “emotion* suppression” OR ruminati* OR distraction |
| 5. | DE “Affect Regulation” OR DE “Emotion Regulation” OR DE “Emotional Regulation” OR DE “Stress and Coping Measures” OR DE “Coping Behavior” OR DE “Coping Style” OR DE “Stress Management” OR DE “Expressive Suppression” OR DE “Emotional Control” OR DE “Anger Control” OR DE “Self-Regulation” OR DE “Cognitive Appraisal” OR DE “Cognitive Control” OR DE “Thought Suppression” OR DE “Suppression (Defense Mechanism)” |
| 6. | S4 OR S5 |
| 7. | “minority stress*” OR discriminat* OR violence OR mistreat* OR oppress* OR marginalize* OR microaggress* OR victimi* OR reject* OR stigma* OR prejudic* OR heterosexis* OR homonegativ* OR harass* OR homophobi* OR transphobi* OR queerphobic*, biphobi*, OR cisgenderis* OR “cis-genderis*” OR cisheterosexis* OR “cis-heterosexis*” OR cisnormativ* OR “cis-normativ*” OR heteronormativ* OR “hetero-normativ*” OR “hypervigilan*” OR “rejection sensitiv*” OR “gender dysphori*” OR “anti-LGBT*” |
| 8. | DE “Minority Stress” OR DE “Marginalization” OR DE “Prejudice” OR DE “Stigma” OR DE “Victimization” OR DE “Discrimination” OR DE “Sex Discrimination” OR DE “Social Discrimination” OR DE “Microaggression” OR DE “Gender Dysphoria” OR DE “Harassment” OR DE “Hate Crimes” OR DE “Hate” OR DE “Aggressive Behavior” OR DE “Oppression” |
| 9. | S7 OR S8 |
| 10. | S3 AND S6 AND S9 |
| 11. | Limited results of S10 to English language and applied the “apply equivalent subjects” expander |

*Note.* Used “All Fields” default option for 3, 6, and 9.

**S4 Table:**
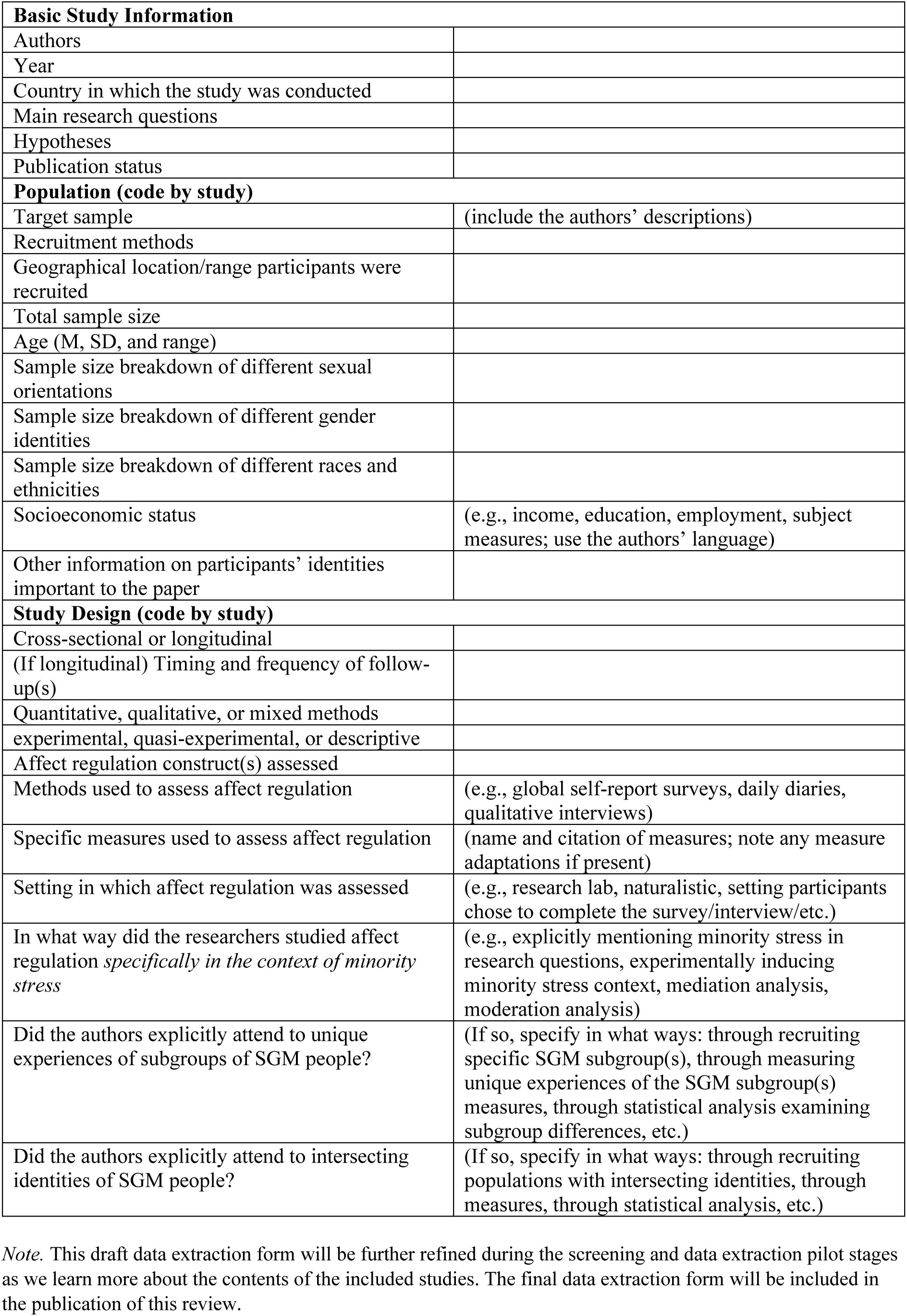
Draft data extraction form.

|  |  |
| --- | --- |
| **Basic Study Information** |  |
| Authors |  |
| Year |  |
| Country in which the study was conducted |  |
| Main research questions |  |
| Hypotheses |  |
| Publication status |  |
| **Population (code by study)** |  |
| Target sample | (include the authors' descriptions) |
| Recruitment methods |  |
| Geographical location/range participants were recruited |  |
| Total sample size |  |
| Age (M, SD, and range) |  |
| Sample size breakdown of different sexual orientations |  |
| Sample size breakdown of different gender identities |  |
| Sample size breakdown of different races and ethnicities |  |
| Socioeconomic status | (e.g., income, education, employment, subject measures; use the authors' language) |
| Other information on participants' identities important to the paper |  |
| **Study Design (code by study)** |  |
| Cross-sectional or longitudinal |  |
| (If longitudinal) Timing and frequency of follow-up(s) |  |
| Quantitative, qualitative, or mixed methods |  |
| experimental, quasi-experimental, or descriptive |  |
| Affect regulation construct(s) assessed |  |
| Methods used to assess affect regulation | (e.g., global self-report surveys, daily diaries, qualitative interviews) |
| Specific measures used to assess affect regulation | (name and citation of measures; note any measure adaptations if present) |
| Setting in which affect regulation was assessed | (e.g., research lab, naturalistic, setting participants chose to complete the survey/interview/etc.) |
| In what way did the researchers studied affect regulation *specifically in the context of minority stress* | (e.g., explicitly mentioning minority stress in research questions, experimentally inducing minority stress context, mediation analysis, moderation analysis) |
| Did the authors explicitly attend to unique experiences of subgroups of SGM people? | (If so, specify in what ways: through recruiting specific SGM subgroup(s), through measuring unique experiences of the SGM subgroup(s) measures, through statistical analysis examining subgroup differences, etc.) |
| Did the authors explicitly attend to intersecting identities of SGM people? | (If so, specify in what ways: through recruiting populations with intersecting identities, through measures, through statistical analysis, etc.) |

*Note.* This draft data extraction form will be further refined during the screening and data extraction pilot stages as we learn more about the contents of the included studies. The final data extraction form will be included in the publication of this review.

